# A nationwide prospective randomized trial for diagnosing developmental disorders demonstrates genome sequencing outperforms standard of care

**DOI:** 10.1101/2025.10.18.25328208

**Authors:** Geysens Mathilde, Souche Erika, Baroni Maria Chiara, Benoit Valérie, Beyltjens Tessi, Bossuyt Wouter, Bours Vincent, Crabbé Marian, Croes Didier, De Smet Matthias, Désir Julie, Devriendt Koenraad, Dheedene Annelies, Dimitrov Boyan Ivanov, Janssens Katrien, Kumps Candy, Lumaka Aimé, Menten Björn, Meuwissen Marije, Monestier Olivier, Mortier Geert, Olsen Catharina, Palmeira Leonor, Peeters Hilde, Pölsler Laura, Revencu Nicole, Roland Dominique, Soblet Julie, Sznajer Yves, Taziaux Sylvie, Van Esch Hilde, Vandeweyer Geert, Vantroys Elise, Vilain Catheline, Wiame Elsa, Van Den Bogaert Kris, Vermeesch Joris Robert

**Author notes:** Corresponding authors Prof. Joris Vermeesch, Prof. Kris Van Den Bogaert.

## Abstract

**Background:** exome (ES) or genome (GS) sequencing are recommended as first- or second-tier molecular tests for patients with developmental disorders (DD), but the clinical utility of GS continues to be debated.

**Methods:** This prospective randomized trial involving all Belgian Human Genetics centers compared the standard of care (SoC) - combining ES and chromosomal microarray analysis or shallow GS - with GS for 567 individuals with unexplained DD.

**Results:** The diagnostic yield of GS was 39.8% (113/284) vs 30% for SoC (85/283) (p=0.015), mainly due to an increased detection of single nucleotide variants and indels (+8.7%). GS also enabled the detection of three non-coding (potential) pathogenic variants. Diagnostic yield was higher for females (45.5%, 97/213) compared to males (28.5%, 101/354) (p<0.001). *De novo* variants were found for 23.6% of patients. Analysis of inherited variants in genes associated with autosomal dominant phenotypes contributed more to the diagnostic yield (3.9%) than X-linked variants (1.9%), and to a similar extent as autosomal recessive variants (4.1%).

**Conclusion:** This nationwide study indicates GS outperforms SoC for the diagnosis of patients with DD in a decentralized hospital setting and well-characterized cohort. The results also highlight the importance of evaluating autosomal dominant inherited variants in genomics analyses for DD.

## 1. BACKGROUND

Developmental disorders (DD) are characterized by early-onset impairments in cognitive, motor, and physical development, and are often associated with intellectual disability (ID) and/or congenital anomalies (CA). Globally, CA affect approximately 2% of life births^1^, while ID impact around 3% of the population^2^. The heterogeneity of both clinical presentations and underlying molecular etiologies presents a challenge to early and accurate diagnosis. The advent of massively parallel sequencing technologies, such as exome (ES) and genome (GS) sequencing, has paved the way for simultaneous assessment of large numbers of genes in a single test, and facilitated the identification of previously unrecognized gene-disease associations^3^. With the introduction of ES, the diagnostic yield for patients with DD has increased to approximately 30%^4,5^. Given the impact of a molecular diagnosis on short- and long-term clinical management of patients, potential therapeutic opportunities, and family and reproductive counseling, ES or GS have been recommended for patients with ID/CA, as a first-tier molecular test or a second-tier test after negative copy-number variant (CNV) analysis^6^.

GS is gradually revealing the involvement of non-coding variants in developmental and numerous other disorders^7-9^ prompting questions about the proportion of missed diagnoses when analyses are confined to the exome. Moreover, GS not only elucidates an additional 90 percent of the genome, it also provides more uniform coverage^10^, improving detection of exonic single-nucleotide variants (SNVs)^11^’^12^ as well as CNVs and structural variants (SVs)^13^. Aside from the pursuit of improving diagnostic yield, GS could act as a potential single molecular test replacing the combination of ES with chromosomal microarray analyses (CMA), shallow GS (sGS) and other technologies, diminishing the burden on the diagnostic laboratories. Schobers *et al*.^14^ evaluated GS’s performance for the detection of thousands of variants identified by various technologies, such as Sanger sequencing, karyotyping, multiplex ligation-dependent probe amplification, CMA, fluorescence in situ hybridization, and ES, and demonstrated non-inferiority for the capture of 95% of variants, with mosaic variants and variants in homologous regions remaining important technical constraints. The streamlining of a sequential diagnostic approach into a single methodology is also expected to have a favorable impact on the time to diagnose, which is crucial in critically ill patients and neonates. The added diagnostic value of GS and its cost-effectiveness, however, remain debated^15,16^.

A meta-analysis of 37 studies, involving 20,068 probands and published between 2011 and 2017, did not observe a significant difference in diagnostic yield between ES and GS^17^. Subsequent studies involving patients with DD assessed GS in cohorts of 32, 38, 52, and 108 undiagnosed individuals following ES, demonstrating increases in diagnostic yield ranging from 3% to 19%^15,18-20^. When offering 150 individuals with neurodevelopmental disorders ES and GS in parallel, Van der Sanden *et al*.^21^ only identified two certain (1.3%) and four potential (2.7%) diagnoses for which GS was required. A recently published direct comparison between GS and SoC molecular tests including ES, observed a difference in diagnostic yield of 3.3% in a cohort of 416 index patients with rare diseases^22^. However, in a broader but also phenotypically diverse cohort of 744 individuals with prior inconclusive ES results, a (likely) pathogenic variant that was only identifiable by GS was detected in 6.3% of cases, increasing to 8.2% when including potential diagnoses^23^.

Thus far, only a single randomized controlled trial (RCT) - the NSIGHT2 trial -systematically compared ES with GS. That study aimed to evaluate rapid sequencing in a cohort of 213 critically ill infants and showed similar diagnostic yields (19% vs 20%) in both study arms^24^. Another RCT compared GS and standard of care (SoC) methodologies without observing significant differences in a cohort of 202 individuals undergoing genetic testing for a broad spectrum of diseases. However, the SoC approaches varied considerably, and ES was only performed for a subset of participants^25^. The intra- and inter-study heterogeneity in included phenotypes, along with variability in applied SoC methodologies and GS analyses -such as including or not the assessment of non-coding regions and SVs-likely contributes to the wide variability observed in study outcomes. Few studies and none of both RCTs specifically assessed individuals with DD. Additionally, most comparative studies between GS and SoC have relied on centralized reference laboratories for GS, which may not accurately reflect its diagnostic performance in decentralized routine clinical settings. In this prospective RCT, we compared the diagnostic yield of the SoC — combining ES and CMA/sGS — with that of GS in a well-characterized cohort of 567 individuals with DD in a nationwide, multicenter, and hospital-based setting.

## 2. MATERIALS AND METHODS

### Patient recruitment and randomization

All eight centers for human genetics in Belgium engaged in this prospective RCT-the BeSolveRD trial - comparing the use of SoC and GS for the molecular diagnosis of DD including ID and CA. The study was registered (ClinicalTrials.gov NCT07051213), approved by the Ethical Commission of UZ Leuven (S64603) and was performed in accordance with the Declaration of Helsinki. A total of 567 participants were recruited between June 2021 and June 2024. Informed consent was obtained from all participants or their legal representatives. Inclusion criteria were (1) moderate to profound ID, (2) mild to moderate ID with recurrence among siblings and parents with a normal intellect, (3) mild to moderate ID and dysmorphism, (4) mild to profound ID and one or more CA, (5) one major CA and dysmorphism and, (6) multiple major CA in two or more different organ systems. CA are considered major when they have a significant impact on the health of the child, whereas rare morphological features with a limited functional or esthetic impact are considered as minor anomalies^26^. Dysmorphism was defined as the presence of at least 3 minor anomalies. Participants were included without age-related restrictions. Probands with a suspicion of an acquired cause, e.g. congenital infection and toxic exposure, were excluded. Probands with prior testing of a gene panel targeting multiple conditions or exome analyses were also excluded. Prior CMA or shallow GS (sGS) was not an exclusion criterion. Participants were recruited by clinical geneticists to ensure detailed phenotypic assessments were performed and thorough family histories were collected, both being essential for variant interpretation. Blood was collected from probands and their parents to enable trio analyses with inheritance based-variant filtering. For 16 probands only one parent was available, no singletons were included. Nineteen pairs of siblings with a similar DD and their parents were also included. Participants were randomized using a blocked randomization list^27^ into the SoC arm of the study (n=283), or the GS arm (n=284) (Figure 1). Data collection and study management were conducted using REDCap^28^.

**Figure 1.**
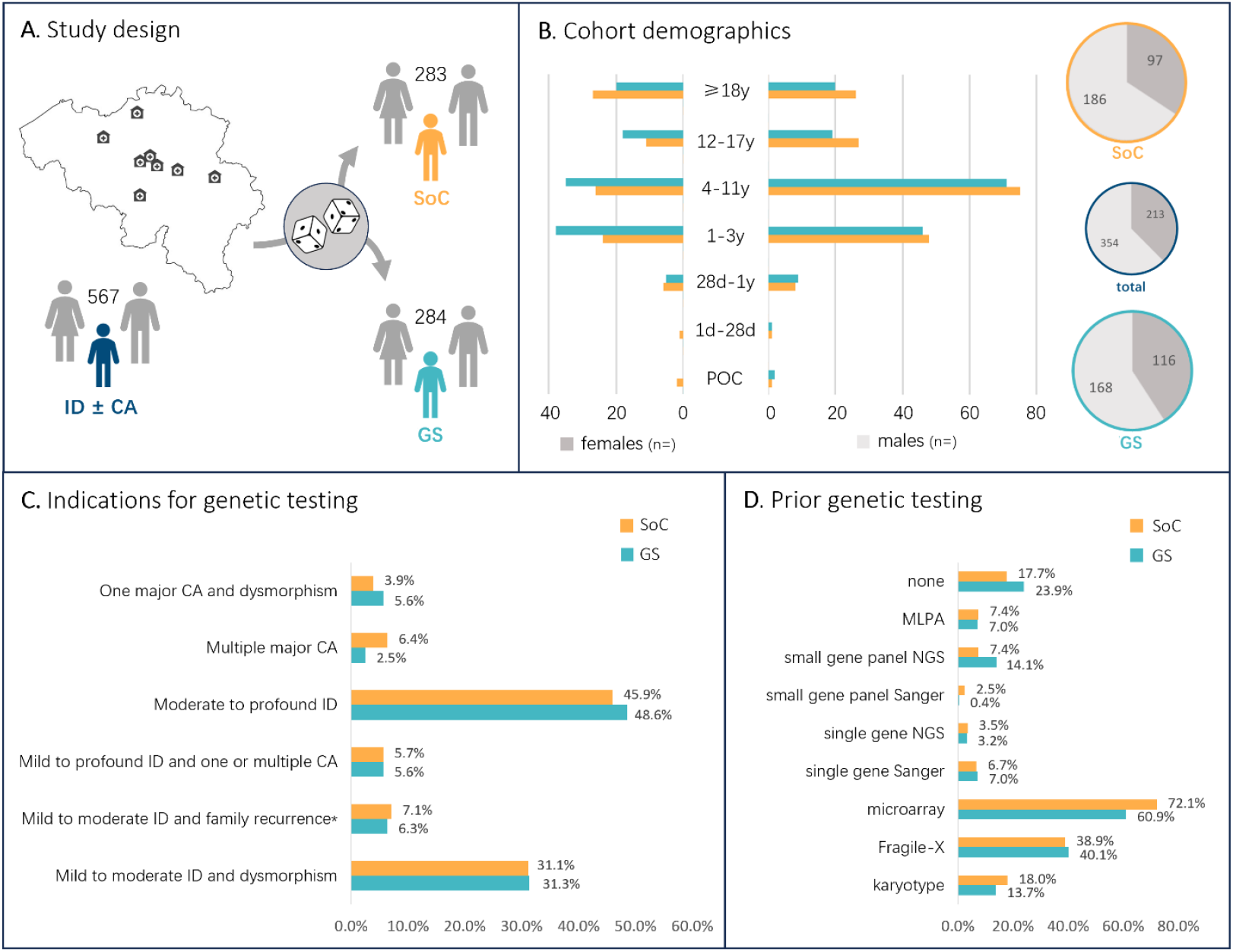
Study design and cohort description. **A**. Illustration of the national multicenter setting of the BeSolveRD study and the randomization of the cohort. **B**. Visualization of the cohort demographics (genetic sex and age) in both study arms. **C**. Inclusion criteria for the study and distribution of the participants across these indication categories for each study arm. **D**. Representation of the different genetic tests performed prior to the study for participants of both study arms. Abbreviations: CA, congenital anomalies; DD, developmental delay; GS, genome sequencing; ID, intellectual disability; POC, products of conception; SoC, standard of care; *parents without ID.

### Sequencing and data processing

In the SoC arm, all probands were investigated using the standard methodology employed at each participating center: CMA- or sGS-based CNV analysis and subsequent ES. ES was performed following the wet lab and bioinformatic protocols that had been previously validated for clinical use by each of the 8 centers (Additional Table 1). The wet lab protocols mainly differed in the shearing methodologies and the exome capture kit used. Differences in bioinformatic processing included the version of the reference genome used (hg19 *vs* hg38 in three and five centers, respectively), preprocessing of the reads prior to mapping (performed in two centers), and base quality score recalibration (conducted in five centers). The tools used for mapping and duplicate marking also differed as did the variant filtering strategies.

**Table 1:** Comparison of diagnostic yields.

| ± AD inherited variants<br>± uncertain diagnoses | GS |  | SoC |  | difference |  |  |
| --- | --- | --- | --- | --- | --- | --- | --- |
|  | total | diagnosed | total | diagnosed | estimate | 95% CI | p-value |
| certain - DN, XL, AR | 284 | 100 | 283 | 76 | 0.084 | [0.007, 0.159] | 0.032 |
| certain - DN, XL, AR, AD | 284 | 113 | 283 | 85 | 0.098 | [0.019, 0.175] | 0.015 |
| certain, uncertain - DN, XL, AR | 284 | 127 | 283 | 102 | 0.087 | [0.006, 0.167] | 0.035 |
| certain, uncertain - DN, XL, AR, AD | 284 | 147 | 283 | 113 | 0.118 | [0.036, 0.199] | 0.005 |

Comparison of the diagnostic yields and statistical significance (according to the Laud<sup>36</sup>, Gart and Nam<sup>37</sup> method) of the estimated difference between both study arms. Abbreviations: AD, autosomal dominant inherited variants in genes associated with dominant phenotypes; AR, autosomal recessive inherited variants; CI, confidence interval; DN, *de novo* variants; GS, Genome Sequencing; SoC, Standard of Care; XL, X-linked inherited variants.

For the GS arm, library preparation and sequencing were performed in five centers for the eight recruiting centers. DNA was sheared using different methods, depending on the site, including sonication (n=2), tagmentation (n=2) and enzymatic fragmentation (n=1). Libraries were prepared without PCR amplification using three distinct kits: Roche KAPA HyperPrep PCR-free (n=2), Illumina DNA PCR-free (n=2), or NEBNext Ultra II FS DNA PCR-free (n=1). Paired-end sequencing was performed on Illumina NovaSeq6000 platforms. Bioinformatic processing was done by each of the eight recruiting centers using a custom pipeline consisting of mapping to the reference genome, duplicate marking and variant calling. Major differences in the analysis pipelines included the version of the reference genome (hg19 *vs* hg38 in two and six centers, respectively), the pre-processing of the reads prior to mapping (done in one center) and the base quality score recalibration (done in five centers). The tools used for duplicate marking and base quality score recalibration also differed. Six of eight centers extracted an exome VCF file prior to interpretation. All centers but two called CNVs from GS data. A mean informative coverage of 42X (+/- 10%) computed by either Picard^29^ or mosdepth^30^ was targeted. Uniformized re-analysis of the data of GS negative samples, including non-coding regions, was performed across the eight centers with the Emedgene software v.32.0.29 (Illumina Inc., San Diego, CA, USA)^31^. Beyond SNVs/indels, the software allows the detection of short tandem repeats (STRs) and SVs.

### Multicenter implementation and cross-validation of GS

SoC methods were validated prior to the study. To ensure a swift implementation and validation of GS in all participating centers, all centers engaged in an inter-laboratory ring trial before patient recruitment. Two Genome-In-A-Bottle (GIAB) samples^32^, HG001 and HG002, were sequenced in the five sequencing centers. Data was shared with the eight analyzing centers so that each center could analyze the samples with its own pipeline and compute coverage statistics, precision, and recall with a standardized protocol. Comparison of the results helped each center pinpoint problematic steps in GS library preparation and sequencing, highlighting the need for large insert sizes and optimal pooling of the libraries to avoid optical duplicates and obtain the expected mean sequencing depth of all samples. The use of different bioinformatic analysis pipelines resulted in only minor differences in the performance of SNV and indel calling as similar precision (HG001: 0.9669 (+/-0.0229); HG002 0.9873 (+/-0.0076)) and recall (HG001: 0.9841 (+/-0.0067); HG002 0.9898 (+/-0.0074)) were obtained in all centers (Additional Figure 1).

### Data analysis and variant interpretation

In the SoC arm, variant interpretation was performed within the accredited diagnostic laboratories of each participating center using their established software platforms and filtering pipelines (Table S1). Identified variants were prioritized based on a gene panel; in three of the eight centers additional exome-wide filtering based on the phenotype of the patient using Human Phenotype Ontology (HPO) terms was applied. In all centers, the variants were further assessed according to Mendelian inheritance modes, starting with *de novo* variants and further investigating X-linked (XL) variants as well as variants that might lead to autosomal recessive (AR) conditions (homozygous and compound heterozygous variants). In addition to these standard analyses, potential disease-causing inherited variants in genes associated with autosomal dominant (AD) conditions were also investigated. Variants were classified according to the American College of Medical Genetics and Genomics/Association for Molecular Pathology and/or Association for Clinical Genomic Science classification guidelines^33,34^. (Likely) pathogenic variants (classes 4 and 5) that could at least partially explain the phenotype of the patient were reported and considered as certain diagnoses. VUS were only reported upon high suspicion of association to the patient’s phenotype. Such VUS, as well as (likely) pathogenic variants with an uncertain contribution to the phenotype, mostly susceptibility factors, were considered as uncertain diagnoses.

In the GS arm of the study, variant filtering and interpretation were performed by the investigators of the same diagnostic laboratories to avoid interpretation biases. In the first phase, GS SNVs/indels as well as CNVs (for six centers), were interpreted using the same software, filters and interpretation criteria as in the SoC arm. Additionally, re-analysis of the data of negative GS cases was performed using the Emedgene software^31^ to tackle potential causal repeat expansions, structural and/or intronic variants. The AI-based filtering and interpretation of variants in this software integrates HPO-based phenotypic information to improve variant prioritization^35^. Statistical significance of the observed differences in diagnostic yield was measured using the score method with skewness correction, based on the approaches of Laud, Gart and Nam^36,37^.

## 3. RESULTS

### Study cohort characteristics

A total of 567 probands were recruited into the study, including 353 males and 214 females. The randomization process resulted in a similar distribution of demographic characteristics and clinical indications between the study arms (Figure 1). Males accounted for 65.7% (186/283) of the SoC arm and 59.2% (168/284) of the GS arm. Most probands were either toddlers (1-3 years old; 156/567, 27.5%) or children (4-11 years old; 207/567, 36.5%), while smaller proportions were infants between one month and one year of age (28/567, 4.9%), adolescents (12-17 years old; 75/567, 13.2%), or adults (≥18 years; 93/567, 16.4%). Given the low uptake of the study in neonates (< 1 months old; 3/567, 0.5%) and in the prenatal setting (products of conception; 5/567, 0.9%), the study findings primarily reflect diagnostic yields in probands older than one month. Moderate to profound ID (268/567, 47.3%) and mild to moderate ID in association with dysmorphism (177/567, 31.2%) constituted the two most frequent indications for genetic testing. The four other indications each contributed for 4.4% to 6.7% (25-38 probands) of the overall cohort. For 377/567 probands a CNV analysis had been performed in the past without identifying an underlying causal variant (204/283, 72.1% in the SoC arm and 173/284, 60.9% in the GS arm). Fragile-X syndrome was tested before inclusion for 159/353 males and 65/214 females and a conventional karyotype had been performed for 90 probands. Sanger sequencing had previously been performed to assess a single gene or a disease-targeted panel of genes for respectively 39 and eight individuals, and NGS had been performed to evaluate a single gene or a disease-targeted panel of genes in 19 and 61 individuals, respectively (Figure 1).

### Diagnostic yield in both study arms

A certain molecular diagnosis was established for 76/283 (26.9%) probands analyzed with SoC methodologies (evaluating *de novo*, XL and AR variants). Most of these variants (70/76, 92.1%) were SNVs or indels identified by ES; the remainder were identified using CMA/sGS and included one SV, four SNVs and one aneuploidy (45,X). Uncertain diagnoses were identified in 26/283 (9.1%) of the probands. In the GS arm (evaluating *de novo*, XL, and AR variants), a certain molecular diagnosis was made for 100/284 probands (35.2%). Similarly as in the SoC arm, the majority (92/100, 92.0%) were SNVs or indels. Seven variants were CNVs and one STR expansion was identified. In addition, uncertain diagnoses (variants of unknown significance and/or unknown contribution to the phenotype) were identified for 27/284 (9.5%) of probands (Figure 2, Additional Figure 2, Additional Table 2).

**Figure 2.**
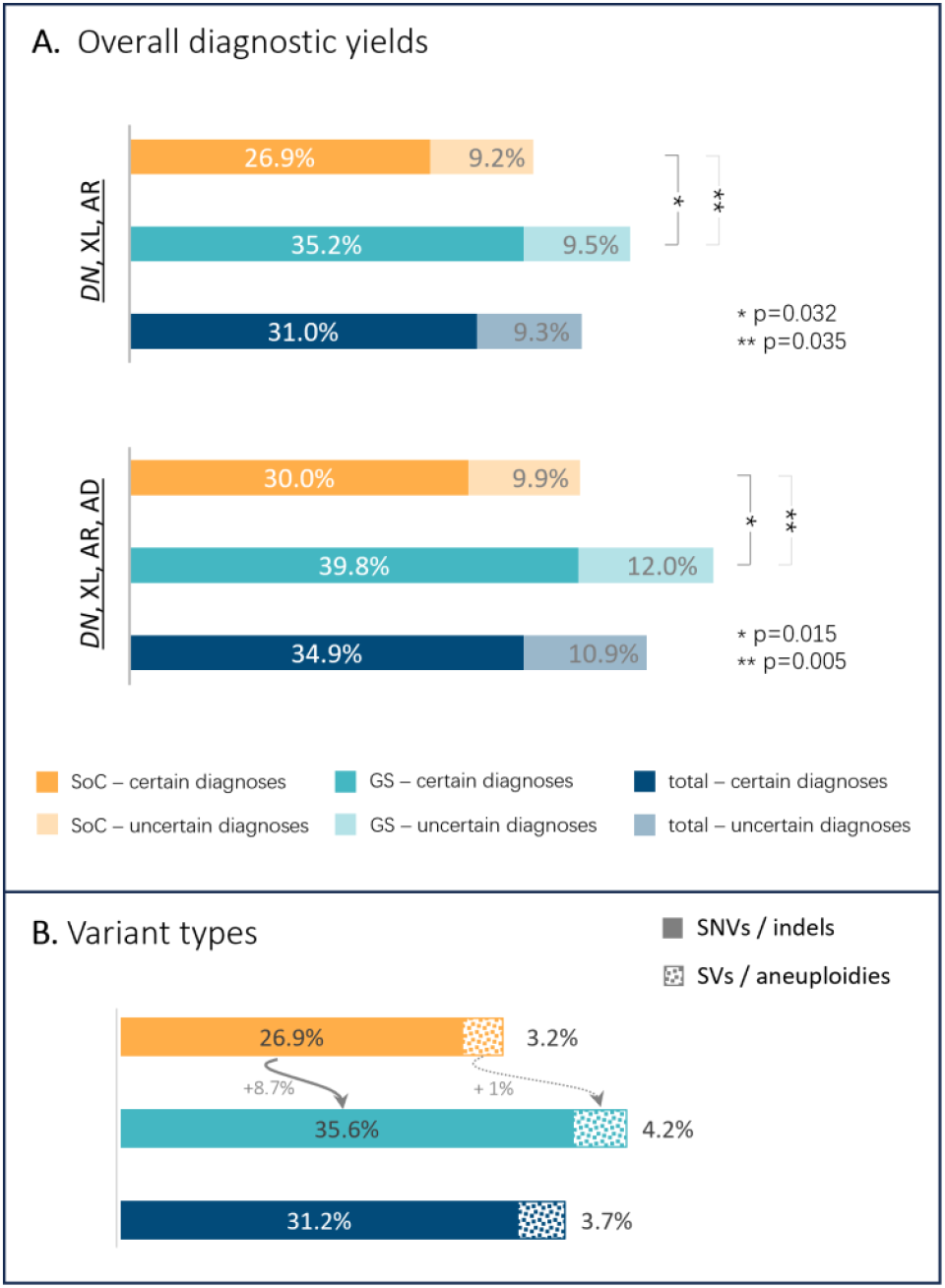
Diagnostic yield in both study arms. **A**. Visualization and comparison of the overall diagnostic yield in both study arms. Dark colors represent certain diagnoses (classes 4 and 5 variants, partially or fully explaining the phenotype), lighter colors represent uncertain diagnoses (class 3 variants and variants with uncertain contribution to the phenotype). **B**. Illustration of the contribution of SNVs/indels *vs* SVs/aneuploidies to the diagnostic yield in each study arm. Statistical significance of the observed differences in diagnostic yield are measured using the score method with skewness correction, based on the approaches of Laud, Gart and Nam^36,37^. Abbreviations: AD, autosomal dominant inherited variants in genes associated with dominant phenotypes; AR, autosomal recessive inherited variants; DN, *de novo* variants; SNV, single nucleotide variant; SoC; standard of care; SV, structural variant; GS, genome sequencing. XL, X-linked inherited variants.

Eight of the certain diagnoses in the GS arm were detected during the systematic reanalysis using the Emedgene platform. One of these was a *de novo* 5’UTR SNV in *MEF2C* (MIM: 600662) NM_002397.5:c.-26C>T. This non-coding variant has been shown to create an upstream new start codon which leads to the translation of an elongated protein^8^. In another proband, a STR expansion in the promotor region of *FMR1* (MIM: 309550), responsible for Fragile-X syndrome, was identified. In two other, unrelated probands, a pathogenic NM_033517.1:c.3679dupG p.(Alal227Glyfs*69) in *SHANK3* (MIM: 606230) had been filtered out in the routine analysis because of its frequency in the GnomAD and exome Sequencing Project population databases, which might be inflated due to sequencing errors as the insertion is affecting a stretch of eight guanines^38^. A fifth variant was NM_145686.4:c.572C>A p.(Thr191Asn) in *MAP4K4* (MIM: 604666). This variant was initially classified as a VUS but reclassified as pathogenic during Emedgene reanalysis due to novel evidence^39^. Three additional diagnoses were uncovered because the applied gene panel did not include the involved genes *(INTS11* (MIM: 611354) NM_0178716:c.1003C>T (p.Gln335Ter), C.1313OT (p.Pro438Leu); *NKX6-2* (MIM: 605955) NM_177400:c.134delTinsGG (p.Leu45ArgfsTer415); *H3F3A* (MIM: 601128) NM_002107:c.365C>T (p.Pro122Leu)).

### Analysis of inherited variants

Beyond the standard analysis of *de novo*, XL, and AR variants, an additional assessment was performed to identify inherited variants in genes associated with AD conditions. This analysis identified nine inherited (likely) pathogenic variants in the SoC and 13 in the GS arm. These 22 additional diagnoses represent 11% of all certain diagnoses and an increased diagnostic yield of 3.9% (Figure 3). For most of these families (18/22) there was no prior suspicion of a dominantly inherited disorder. Seventeen of the 22 variants (77.3%) were interpreted as fully explaining the patient’s phenotype while five were recurrent CNVs considered risk factors for ID and were thought to partially explain the clinical presentation of the patient. As a comparison, the *de novo* analyses enabled identification of a (likely) pathogenic variant for 134 patients (23.6%) while the XL analysis was contributive for only 11 patients (1.9%) and the recessive analysis for 23 patients (4.1%). AD inherited variants of unknown significance and/or unknown contribution to the phenotype were retained in another 13 patients (2.3%) (Additional Figure 3). One of these variants was a ∼30kb deletion that includes the *5’UTR* first exon of *ANKRD11* (MIM: 611192). Deletions of this region have been associated with downregulation of *ANKRDll* transcription, resulting in KBG syndrome^40^.

**Figure 3.**
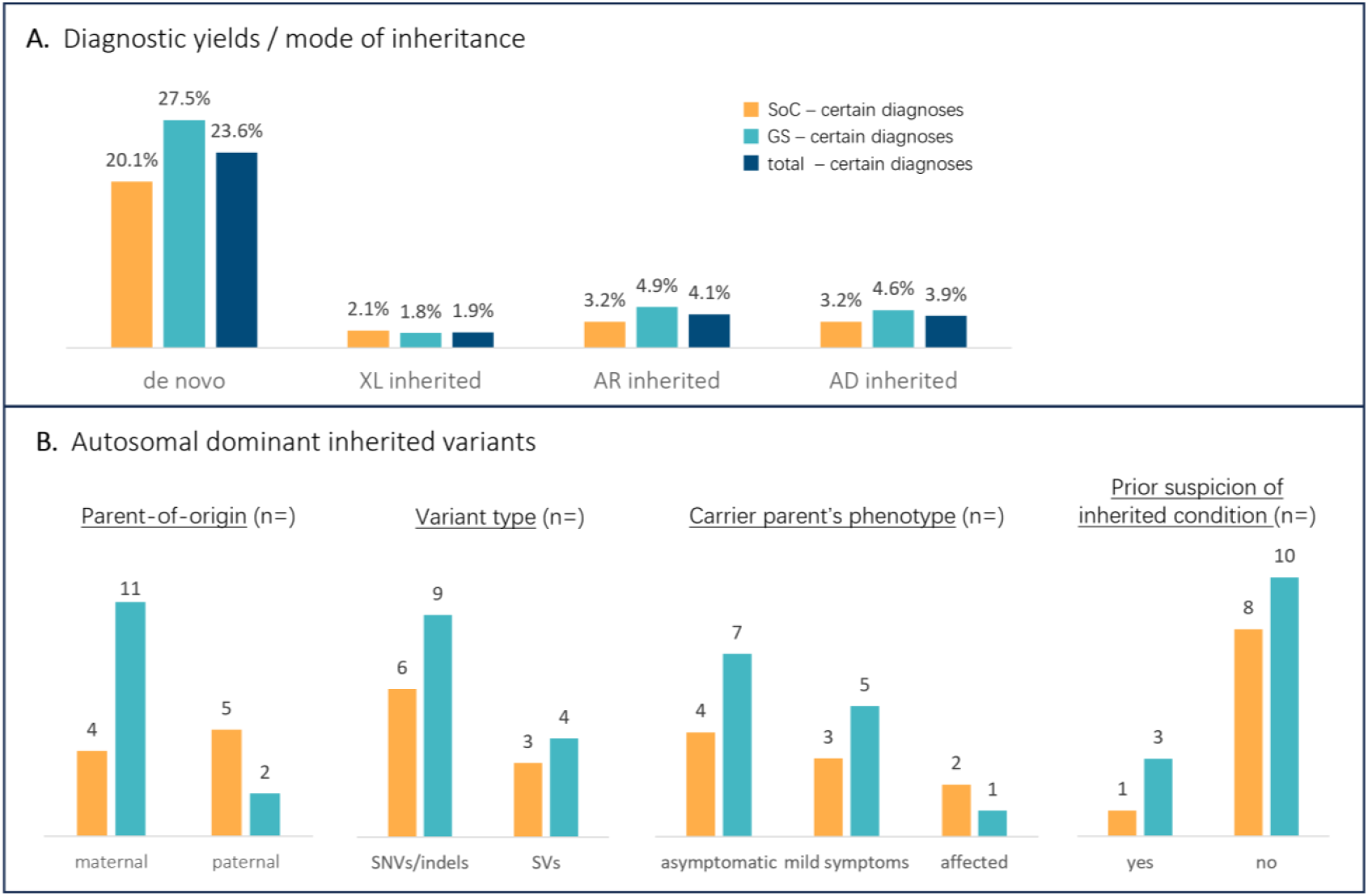
Autosomal dominant inherited variants. **A**. Comparison of the diagnostic yield of the different inheritance modes analyses (bar chart) and contribution (%) of each inheritance analysis to the overall diagnostic yield (pie chart). **B**. Visualization of the parental origin of AD inherited variants, the type of AD inherited variants as well as the clinical context (parental phenotype) in which they were identified. Abbreviations: AD, autosomal dominant inherited variant in genes associated with dominant phenotypes; AR, autosomal recessive inherited variants; DN, *de novo* variants; XL, X-linked inherited variants.

The variant allele frequency (VAF) of the inherited SNVs (0.30 to 0.57) was consistent with heterozygous carriership for most (20/22) carrier parents (Additional Table 3). Two causal variants were likely inherited from a mosaic parent (VAF <0.25), which is a plausible explanation for the absence of a phenotype in the parent. Another cause of differences in parental and patient phenotypes is imprinting, which are involved in the variant disrupting the *GNAS* (MIM: 139320) gene^41^. Overall, inheritance was maternal for 15/22 cases (68.2%) and paternal for 7/22 (31.8%). Half of the carrier parents (11/22) were reported to be asymptomatic. In eight, mild symptoms of the disease could be identified, and three presented more severe symptoms. For all genes in which (likely) pathogenic inherited variants were identified, variable expression and reduced penetrance has been described in the literature.

### Added value of genome sequencing

Comparing the diagnostic yield in both arms of the study (evaluating *de novo*, XL and AR variants), we observed a significant increased yield of 8.4% (95% CI: 0.7% - 15.9%, p=0.032) in the GS arm (SoC 26.9% (76/283), GS 35.2% (100/284)). After analysis of inherited AD variants, the added value of GS increased to 9.8% (95% CI: 1.9% - 17.5%, p=0.015) (Table 1), which corresponds to a relative added diagnostic yield of 32.7%.

The impact of SVs/aneuploidies on the yield difference was minor (+1.0%), with the number of (likely) pathogenic SVs being low in both arms (seven CNVs and one SV in the SoC arm and 11 CNVs in the GS arm). The added diagnostic yield in the GS arm primarily reflects an increase in the detection of SNVs/indels, from 26.9% in the SoC arm to 35.6% in the GS arm (+8.7%, 95% CI: 0.011 - 0.163, p=0.025). Of the SNVs/indels detected in the GS arm, only one variant was not disrupting the coding genome or splice sites, namely the 5’UTR NM_002397.5:c.-26C>T variant in *MEF2C* (MIM: 600662). We investigated whether some of the SNVs/indels identified with GS were located in exonic regions poorly covered in the ES data, but the mean informative coverage of the genomic loci of all GS-detected SNV/indels in the exome data was >20x (Additional Table 4). When comparing the yield for each variant subtype, we observed a significant difference in the number of out-of-frame indels, with 26 frameshift variants identified in the GS arm in contrast to 10 in the SoC arm (Figure 4). This difference corresponds to an additional 5.6% (95% CI: 1.7% - 9,8%, p=0.006) of patients for whom an out-of-frame indel was identified in the GS arm. GS also enabled diagnosis of one proband with Fragile-X syndrome, one of the most prevalent causes of ID in males^42^, which is caused by a tandem repeat expansion in the *FMR1* (MIM: 309550) promotor region. Among the CNVs, GS identified a small *5’UTR* deletion potentially downregulating *ANKRD11* (MIM: 611192),a VUS which would have been missed using SoC.

**Figure 4.**
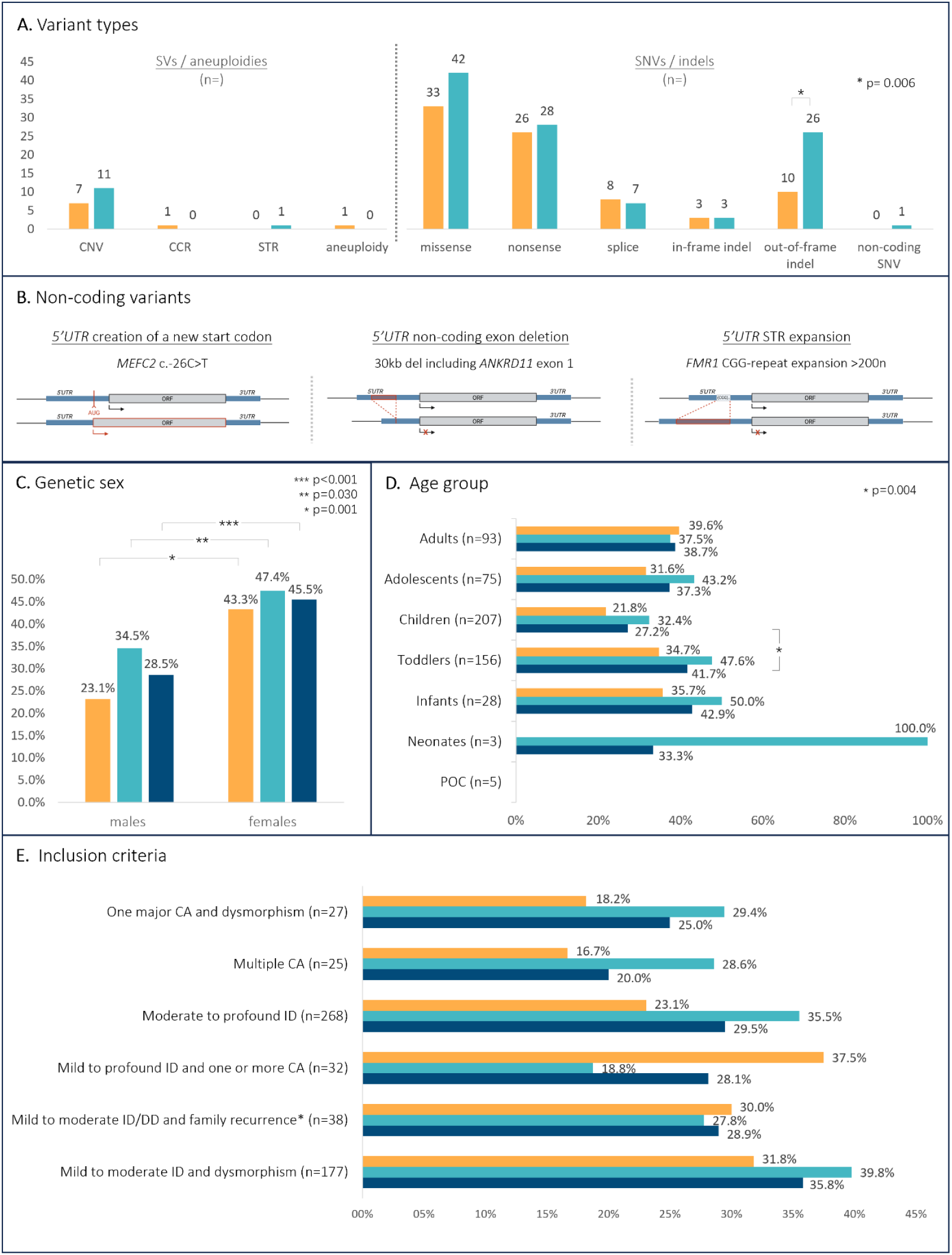
Variant types and demographic subcategories. **A**. Comparison of the number of variants identified in each study arm for each variant type (n=) **B**. Illustration of the three non-coding variants identified by GS. **C**. Illustration of the difference in diagnostic yield between males and females for each study arm as well as the whole cohort. **D.-E**. Graphical representations of the diagnostic yield for different age groups (**D**.) and indication categories **(E.)**. Statistical significance of the observed differences in diagnostic yield are measured using the score method with skewness correction, based on the approaches of Laud, Gart and Nam^36,37^. Abbreviations: CA, congenital anomaly; CCR, complex chromosomal rearrangement; CNV, copy number variant; DD, developmental delay; GS, genome sequencing; ID, intellectual disability; PoC, products of conception; SNV, single nucleotide variant; STR, short tandem repeat; SV, structural variant; UTR, untranslated region.

We further assessed whether certain analytical strategies or characteristics of the cohorts in each of the arms could have impacted diagnostic yields. We reviewed the distribution of the 19 sibling pairs across both study arms and evaluated its potential impact on the results. Excluding one sibling from each pair when both had the same diagnosis and were randomized to the same study arm had only a minor effect on overall outcomes, with diagnostic yields of 30.0% (82/273) in the SoC arm and 40.3% (112/278) in the GS arm (diagnostic yield difference of 10.3%, p=0.012). We also investigated the potential influence of expanded gene panels, less stringent population-based filtering, and recent literature evidence in the GS Emedgene reanalysis. Correcting for the six molecular diagnoses for which detection was explained by these parameters, we still measured an increase in diagnostic yield of 8.1% (95% CI: 0.2% -15.9%, p=0.045) in the GS arm.

We then explored whether differences in yield could be influenced by indication categories or age groups distribution. We mainly observed lower yields for the patients for whom CA are reported without associated ID (25% vs an average of 36% for the categories including ID). Comparing the yield across the age groups, the yield was lower in children than in other age groups. The only difference reaching significance among indication categories or age groups, was the difference in yield between toddlers (65/156, 41.7%) and children (56/206, 27.2%) (difference of 14.5%, 95% CI: 4.8% - 24;4%, p=0.004). However, distribution of all subcategories was relatively homogeneous amongst study arms (Figure 1). Comparing the diagnostic yield between males and females, 45.5% (97/213) of females obtained a diagnosis vs 28.5% (101/354) of males (Figures 4, Additional Figure 4, Additional Tables 5&6), corresponding to a significant difference of 17% (95% CI: 8.7% - 31.7%, pcO.OOl). We therefore evaluated to what extent the slight difference in male/female ratio in both study arms might affect our results. After adjusting for this difference, the diagnostic yield gap was estimated at 9.0% (95% CI: 1.3% - 16.6%, p=0.022) (Additional Table 5). Combining the potential influence of the difference in yield between both sexes with the corrections for the Emedgene reanalysis reduces the diagnostic yield gap between GS and SoC to 7.3% (95% CI: -0.6% to 15.1%, p = 0.069).

Finally, we evaluated the potential impact of the distribution of autism, a phenotypic trait associated with lower diagnostic yields^43,44^. In our cohort, the diagnostic yield in probands with reported autism or autistic behavior was 24.1%. Autism or autistic behavior was more frequent among the unsolved (131/306, 42.8%) than the solved cases (45/198, 22.7%) and in males (137/353, 38.8%) than females (54/214, 25.2%), which might explain part of the observed difference in yield between genetic sexes. However, the proportion of probands with autism or autistic behavior was higher in the GS arm (105/284, 37.0%) than the SoC arm (86/283, 30.4%) (Additional Figure 5). Hence, correcting for this feature would increase the diagnostic yield of GS. Among the other frequent phenotypic traits, higher diagnostic yields were observed in probands with hypotonia (23/39, 59.0%), hypertelorism (16/36, 44.4%), microcephaly (24/55, 43.6%), or short stature (19/44, 43.2%) (Additional Figure 6). However, no significant differences were observed in the distribution of the frequent phenotypic traits across both study arms except for autistic features and moderate ID, the latter being associated with a slightly higher, though non-significant, diagnostic yield (31/83, 37.3% vs 198/567, 34.9% overall) and being more common in the SoC arm (50/283, 17.7% vs 33/284, 11.6% in the GS arm). As a result, adjusting for this factor would have marginally increased the yield of GS.

## 4. DISCUSSION

This multicenter, nationwide, RCT implemented clinical-grade GS, demonstrating its feasibility in a decentralized healthcare system and evaluating its diagnostic utility in patients with DD. A certain molecular diagnosis was made in 39.8% of probands investigated with GS, compared with 30% of probands evaluated using a combination of ES with CMA/sGS (SoC). Uncertain diagnoses were identified in an additional 12.3% and 9.5% of probands in the GS and SoC arms, respectively. Our findings indicate a significant 9.8% (95% CI: 1.9% - 17.5%, p=0.015) increase in diagnostic yield with GS. This observed benefit slightly exceeds the 8.2% added value reported in a recent large study assessing GS utility in patients with prior non-contributory ES^48^. These observations contrast with earlier studies that generally reported smaller and non-significant differences, but might have been limited by limited sample sizes, heterogeneous patients’ cohorts, and variability in SoC protocols^18,25^.

Most (likely) pathogenic variants identified in this trial were exonic SNVs/indels, although a slight increase in the detection of disease-causing SVs and three non-coding variants were observed in the GS arm. The higher yet less uniform coverage of ES has been demonstrated in multiple studies^45,46^and direct comparisons between ES and GS have identified pathogenic coding SNVs/indels missed by ES due to localized coverage drops^11^. Our indirect comparison of ES data did not reveal GS (likely) pathogenic variants in regions poorly covered by ES. As exome capture probes are designed based on the reference genome, we hypothesize that variants with lower sequence homology to the reference may be less effectively captured, leading to reduced representation in the sequencing data. Dissection of the variant types mainly showed a significant increase in the detection of indels in this study.

While the detection rate of CNVs in present cohort is low, we expect these results to be an underestimation of the CNV yield that would be achieved when applying GS as a first-tier genetic test in the future. Over half the cohort (377/567, 66.5%) indeed consisted of probands with prior negative CMA/sGS-based CNV analysis, and pathogenic CNVs are identified in approximately 12% of patients undergoing first-tier CMA/sGS for ID/CA^47^’^48^. Additionally, while GS has been reported to improve CNV calling^49^’^50^, we only identified one candidate disease-causing CNV that would have been missed by SoC, a *5’UTR* deletion potentially disrupting *ANKRD11* (MIM: 611192). In this cohort, GS identified one causal STR expansion in the *FMR1* (MIM: 309550) promotor region, which could not have been detected by SoC. In contrast, in a GS cohort of 744 families, Wojcik *et al*.^23^ detected six STRs and eight inversions or complex rearrangements. The higher number of STR expansions is likely due to differences in the patient population, which included individuals with ataxia and neuromuscular disorders, while the difference in detection of SVs likely reflects the difference in software used for SV detection, as our routine and Emedgene analyses of GS data did not include tools designed for detecting inversions and complex rearrangements.

Beyond the intrinsic technical advantages of GS, most notably its coverage width and uniformity, the combination of GS with more recent analytical software might explain part of the observed difference in the present study. In addition to advances in bioinformatic pipelines, the integration of AI-driven variant prioritization tools allows for less stringent variant filtering strategies while preserving efficiency in the variant interpretation phase. In the present cohort, five of 113 GS diagnoses benefited from the expanded analytical approach through Emedgene. We therefore anticipate that implementing similar analytical and interpretation tools within the SoC could help reduce part of the diagnostic gap between ES and GS. As short-read mapping and SV calling algorithms continuously improve - integrating graph-based genome representations, multigenome mapping, and machine learning based variant detection^51^ - diagnostic yields are expected to further increase, potentially widening the gap between ES and GS through enhanced detection of inversion, complex rearrangements and other SVs.

Furthermore, in addition to increasing the diagnostic yield, GS streamlines the diagnostic process by enabling concurrent detection of SNVs/indels, CNVs, SVs, and STRs^14^. Replacing a sequential diagnostic process with a single test not only enables rapid comprehensive genetic testing^52^, it also reduces the amount of DNA required, simplifying validation processes and lowering costs^16^. Another advantage of GS is the potential to reanalyze the sequencing data as knowledge and insights into the non-coding genome continue to expand^53^. For example, post hoc targeted reanalysis of the *RNU4-2* small nuclear RNA gene, which was recently identified as the cause of 0.5% of neurodevelopmental disorders, resulted in one additional diagnosis among our inconclusive GS samples^54,55^.

Despite the advantages of short-read GS, certain limitations persist. Low-complexity or repetitive regions^56^ as well as certain CNVs, SVs, and STRs^57-59^ remain challenging to resolve using short read GS alone. Epigenetic and mosaic variants represent additional limitations of conventional short-read GS, both of which are increasingly recognized as contributors to DD^60,61^. Long-read sequencing technologies offer complementary potential for the resolution of repetitive regions as well as the detection of complex and epigenetic variants, but are not widely applied in clinical diagnostics yet due to technical and cost-related constraints.

To date, GS studies have mainly been performed in centralized reference or research laboratories. Here, we demonstrate the potential to implement clinical-grade GS inside the diagnostic facilities of the Centers for Human Genetics at a national level, across five sequencing and eight clinical interpretation sites. This local implementation in tertiary care centers is an important factor to consider for patient care, since the diagnostic utility of GS/ES in hospital-based settings has been shown to be qualitatively higher than in external reference laboratories^17^.

Most clinical laboratories focus on the identification of *de novo*, XL, and AR diseases, while inherited AD variants being usually filtered out when performing trio analyses for sporadic DD. Although case reports^62^ and association studies^63,64^ have highlighted the relevance of AD inherited variants in DD, a systematic evaluation of the impact of assessing these variants is lacking. The results of this study reveal that the frequency of disease-causing AD inherited variants is comparable to that of AR causes of DD in our population and approximately twice as high as that of pathogenic XL variants, arguing in favor of including inherited AD variants in the standard work-up of patients with DD, even when both parents appear asymptomatic. While the interpretation of and counseling related to these variants can be difficult, their identification and reporting remain of major importance for both the probands and their family members given the high recurrence risk and potential need for medical follow-up of undiagnosed family members with a milder phenotype. For example, two maternally inherited pathogenic variants were identified in *PTEN* (MIM: 601728), which is associated with Cowden syndrome, a condition associated with a high risk of malignancy that necessitates lifelong medical follow-up.

Our results also demonstrate a significantly higher diagnostic yield in females compared to males. This trend is observed across all indication categories and remains significant among probands with moderate to profound ID with an estimated yield difference of 19.7% (p=0.001). These findings align with previous ES analyses of 13,500 probands with DD in the UK and Ireland^65^, which showed lower odds of diagnosis in males, and reflect the higher prevalence of rare, high-risk variants observed in females with neurodevelopmental disorders^66^. The difference in diagnostic yield between females (47.5%, 76/160) and males (35.0%, 77/220) remains significant, albeit reduced (estimated yield difference of 12.5%, p=0.014), after excluding probands with autism or autistic features. While further research is needed to fully understand potential biological but also sociological factors underlying this difference, future studies comparing diagnostic yields would benefit from matching cohorts to minimize biases related to sex distribution.

## 5. CONCLUSION

Our results provide compelling evidence for the added value of clinical-grade GS in patients with ID/CA, showing a 9.8% increase in diagnostic yield compared with the current SoC, demonstrating a strong incentive for the implementation of GS as first-tier diagnostic test. We expect a further improvement of the diagnostic yield with growing understanding of non-coding variants and implementation of tools specifically developed for the detection of SVs, especially inversions and complex rearrangements.

## Data Availability

Information regarding the phenotype and molecular diagnosis of study participants are shared in the 
supplementary files. The raw sequencing data generated during this study will be shared on EGA (EGAC00001003120).

## Ethics approval and consent to participate

The study was approved by the Ethical Commission of UZ Leuven (S64603) and was performed in accordance with the Declaration of Helsinki. Informed consent was obtained from all participants or their legal representatives.

## Consent for publication

Consent for publication was obtained from all participants or their legal representatives.

## Availability of data and materials

Information regarding the phenotype and molecular diagnosis of study participants are shared in the supplementary materials. The datasets generated and analyzed during the current study will be available through the EGA repository (EGAC00001003120).

## Competing interests

Illumina Inc. (San Diego, CA, USA) provided the consumables for GS of the samples used for the ring trial and the 284 probands and their parent(s) sequenced in the GS arm, as well as access to the Emedgene platform. The authors declare that they have no other competing interests.

## Funding

Illumina Inc. (San Diego, CA, USA) provided the consumables for GS of the samples used for the ring trial and the 284 probands and their parent(s) sequenced in the GS arm, as well as access to the Emedgene platform. This work was additionally supported by grants from the KU Leuven, C1-C14/22/125 and by FWO-TBM grant T-003819N, FWO grant G0A2622N to VJR.

### Authors’ contribution

Conceptualization: VDBK, VJR - Methodology: SE, BW, LA, BM, MM, OC, Ple, RN, SJ, VDBK - Software: SE, CD, DSM, OC, Ple, VG - Validation: SE, BW, CO, PLe, MB - Formal analysis: GM, SE - Investigations: GM, BMC, BV, DA, JK, LA, MM, MO, RN, SJ, TZ, VE, WE, VDBK - Resources (patient recruitment): GM, BT, BV, DJ, DK, DBI, CK, LA, MM, MG, PH, PLa, RN, SY, VEH, VC - Data curation : GM, SE, CM - Writing (original draft) : GM, SE, VJR - Writing (review and editing) : GM, SE, BMC, BV, CM, DJ, DK, JK, KC, LA, MM, MG, OC, PH, PLe, PLa, RN, RD, SY, VEH, VDBK, VJR - Visualization: GM, SE, CM - Supervision: VDBK, VJR - Project administration: CM, VJR - Funding acquisition : VJR.

## Acknowledgements

We express our gratitude to all patients and families who agreed to participate to the study. We thank D. Zhang and J. Surtihadi for their contribution to the statistical analyses and Raye Alford (Illumina, Inc.) for critical review of the manuscript.

## Notes

### Competing Interest Statement

Illumina Inc. (San Diego, CA, USA) provided the consumables for WGS of the samples used for the ring trial and the 284 probands and their parent(s) sequenced in the WGS arm, as well as access to the Emedgene platform.

### Clinical Trial

NCT07051213

### Funding Statement

Illumina Inc. (San Diego, CA, USA) provided the consumables for WGS of the samples used for the ring trial and the 284 probands and their parent(s) sequenced in the WGS arm, as well as access to the Emedgene platform. This work was additionally supported by grants from the KU Leuven, C1-C14/22/125 and by FWO-TBM grant T-003819N, FWO grant G0A2622N to VJR.

### Author Declarations

The Ethical Commission of the University Hospitals of Leuven (UZ Leuven) gave ethical approval for this work (S64603).

