## Additional files for "A nationwide prospective randomized trial for diagnosing developmental disorders demonstrates genome sequencing outperforms standard of care"

**Additional Figure 1:** Performance of SNVs and indels detection across the 8 genetic centers.

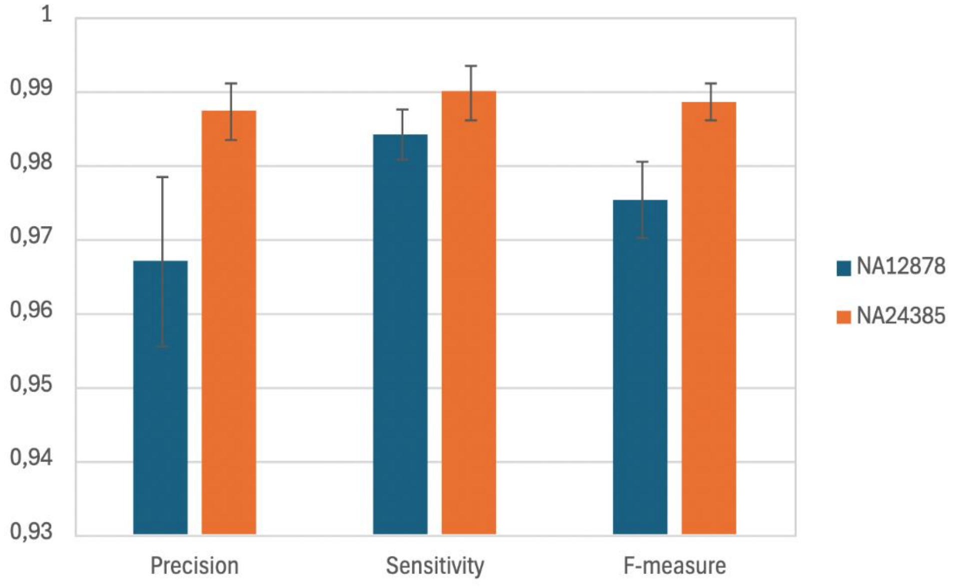


Additional Figure 1. Mean precision, sensitivity and F-measure for the detection of SNVs/indels using GS for the two Genome-In-A-Bottle cell lines, HG001 (=NA12878, in blue) and HG002 (=NA24385, in orange), across the 8 genetic centers. The error bars represent the standard deviation of the values in the different genetic centers.
*Abbreviations:* GS, genome sequencing; SNV, single nucleotide variant.

**Additional figure 2:** Study flow diagram.

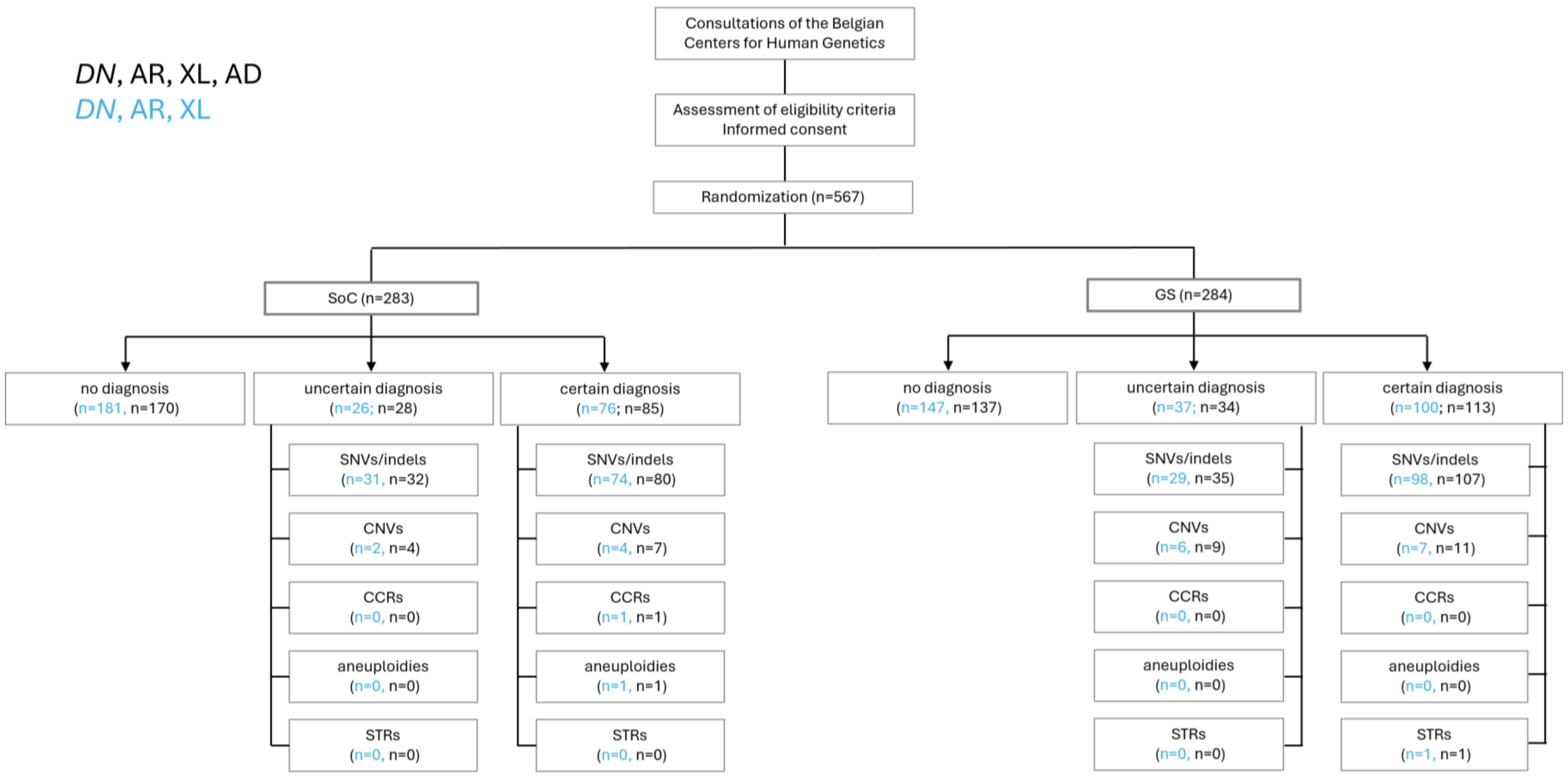


Additional Figure 2. Distribution of included probands in both study arms as well as primary outcome for each individual, being the absence of a diagnosis, an uncertain diagnosis or a certain diagnosis. In addition, the numbers of identified variants are given for each variant subtype. The numbers are given without analysis of AD inherited variants in blue, and after completing this analysis in black.

For the primary outcome, probands in which both a certain diagnosis and an additional uncertain diagnosis were identified, are only accounted in the certain diagnosis group. For the variant types, each identified variant is taken into account. Since multiple variants were identified in some probands, the total amounts of variants may differ from the total amount of probands with a diagnosis.

*Abbreviations:* AD, autosomal dominant inherited; AR, autosomal recessive inherited; DN, *de novo*; CCR, complex chromosomal rearrangement; CNV, copy number variant; GS, genome sequencing; SoC, standard of care; SNV, single nucleotide variant; STR, short tandem repeat; SV, structural variant; XL, X-linked inherited.

**Additional Figure 3 :** Diagnostic yield for each mode of inheritance.

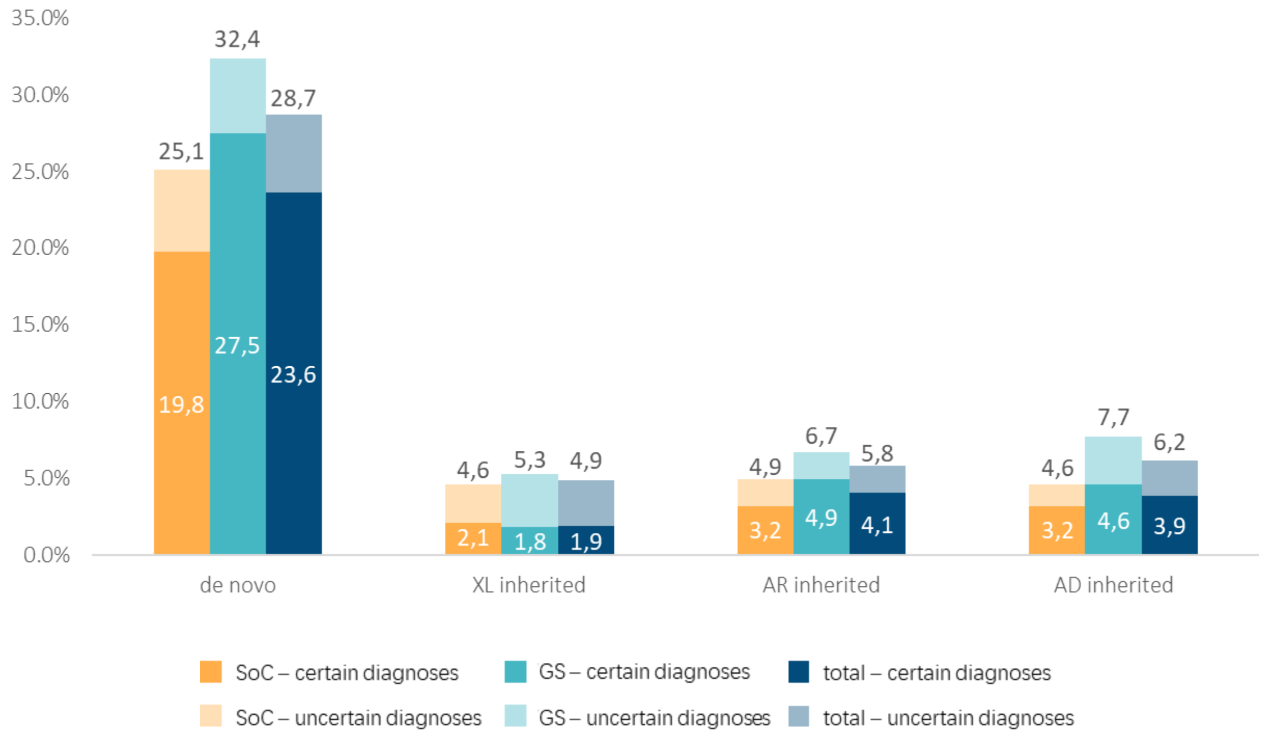


Additional Figure 3. Diagnostic yield for each mode of inheritance, in the SoC arm, the GS arm, and the overall cohort, including uncertain diagnoses. Each bar represents the diagnostic yield of each inheritance analysis, the dark part representing the certain diagnoses as defined in the manuscript (class 4 and 5 variants, with partial or full contribution to the phenotype), whereas the lighter bars represent the uncertain diagnoses (class 3 variants and/or variants with uncertain contribution to the phenotype). Data labels in the dark box give the yield (percentage) for the certain diagnoses, upper data labels give the yield (total percentage) for both certain and uncertain diagnoses.

*Abbreviations:* AD, autosomal dominant inherited; AR, autosomal recessive inherited; GS, genome sequencing; SoC, standard of care; XL, X-linked inherited.

**Additional Figure 4 :** Diagnostic yield for each genetic sex.

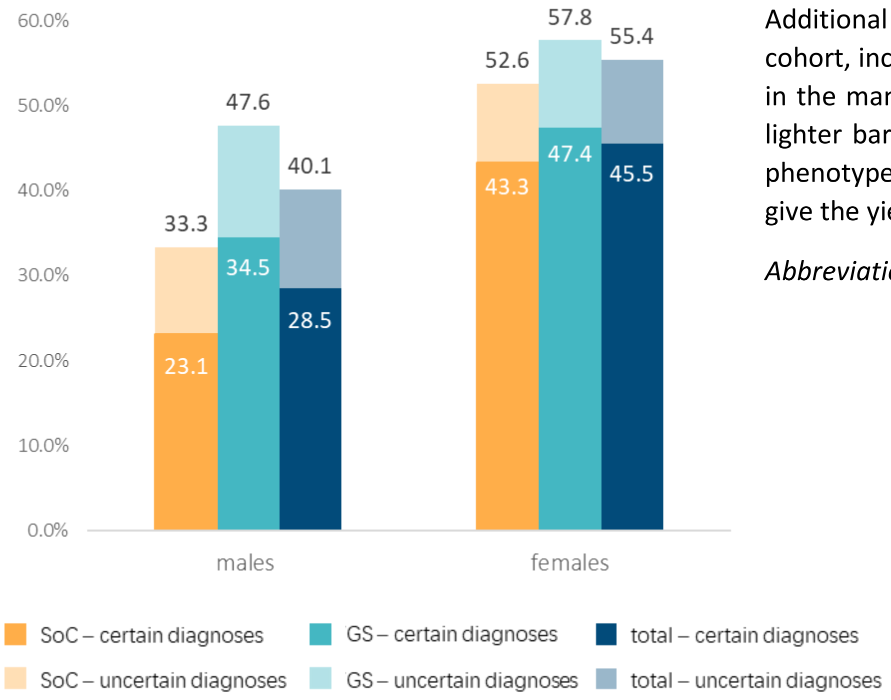


Additional Figure 4. Diagnostic yields in males vs females, in the SoC arm, the GS arm, and over the overall cohort, including uncertain diagnoses. The dark part of each bar represents the certain diagnoses as defined in the manuscript (class 4 and 5 variants, with partial or full contribution to the phenotype), whereas the lighter bars represent the uncertain diagnoses (class 3 and/or variants with uncertain contribution to the phenotype). Data labels in the dark box give the yield (percentage) for the certain diagnoses, upper data labels give the yield (total percentage) for both certain and uncertain diagnoses.

*Abbreviations:* GS, genome sequencing; SoC, standard of care.

**Additional Figure 5 :** Autism and autistic behavior.

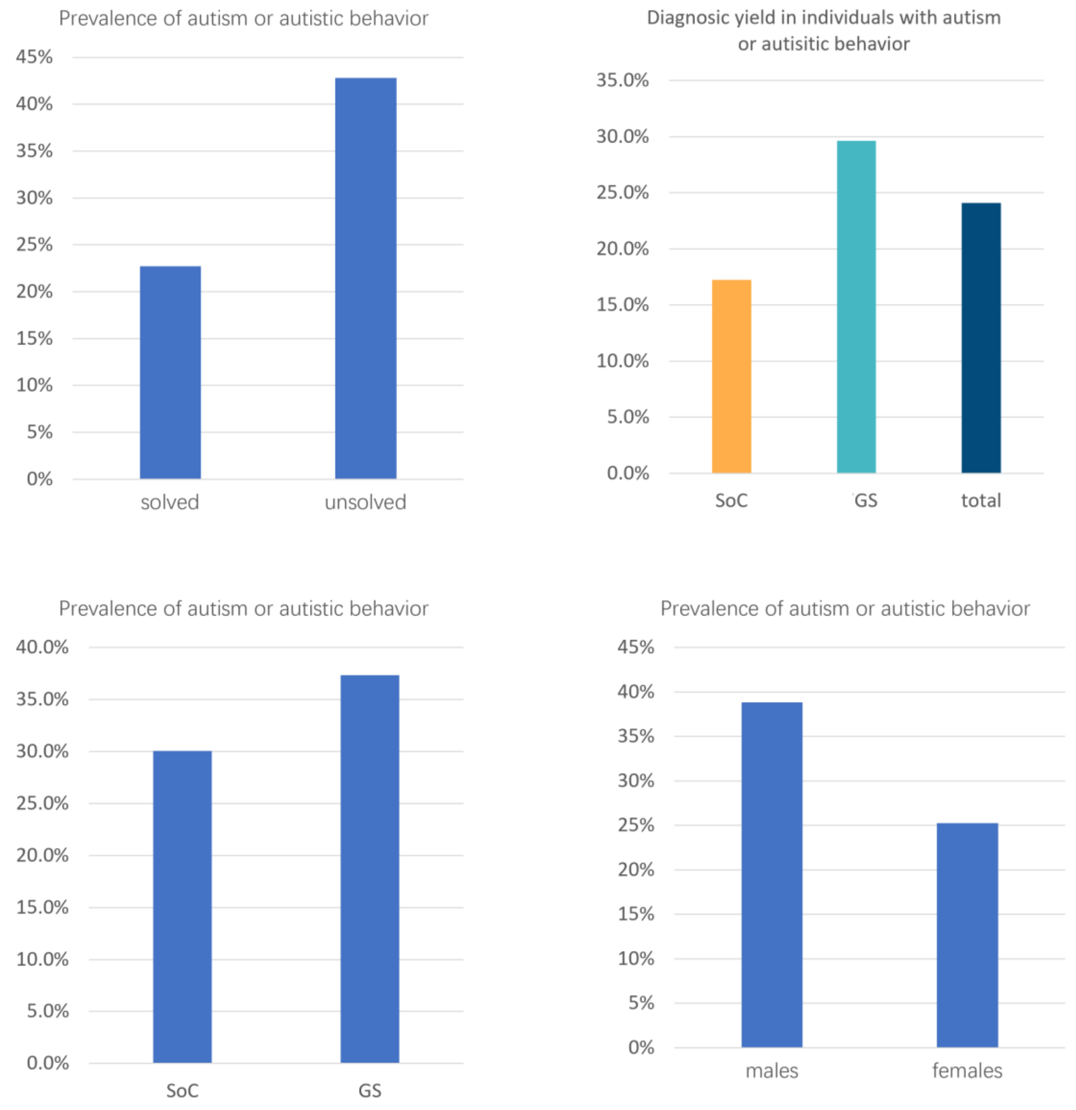


Additional Figure 5. Comparison of autism and autistic behavior prevalence across different groups: solved vs unsolved participants (top left), Soc vs GS study arms (bottom left), and males vs females (bottom right). The diagnostic yield among probands with reported autism or autistic behavior is shown in the top right panel.

*Abbreviations:* GS, genome sequencing; SoC, standard of care.

**Additional Figure 6 :** Diagnostic yield for the 20 most prevalent HPO terms.

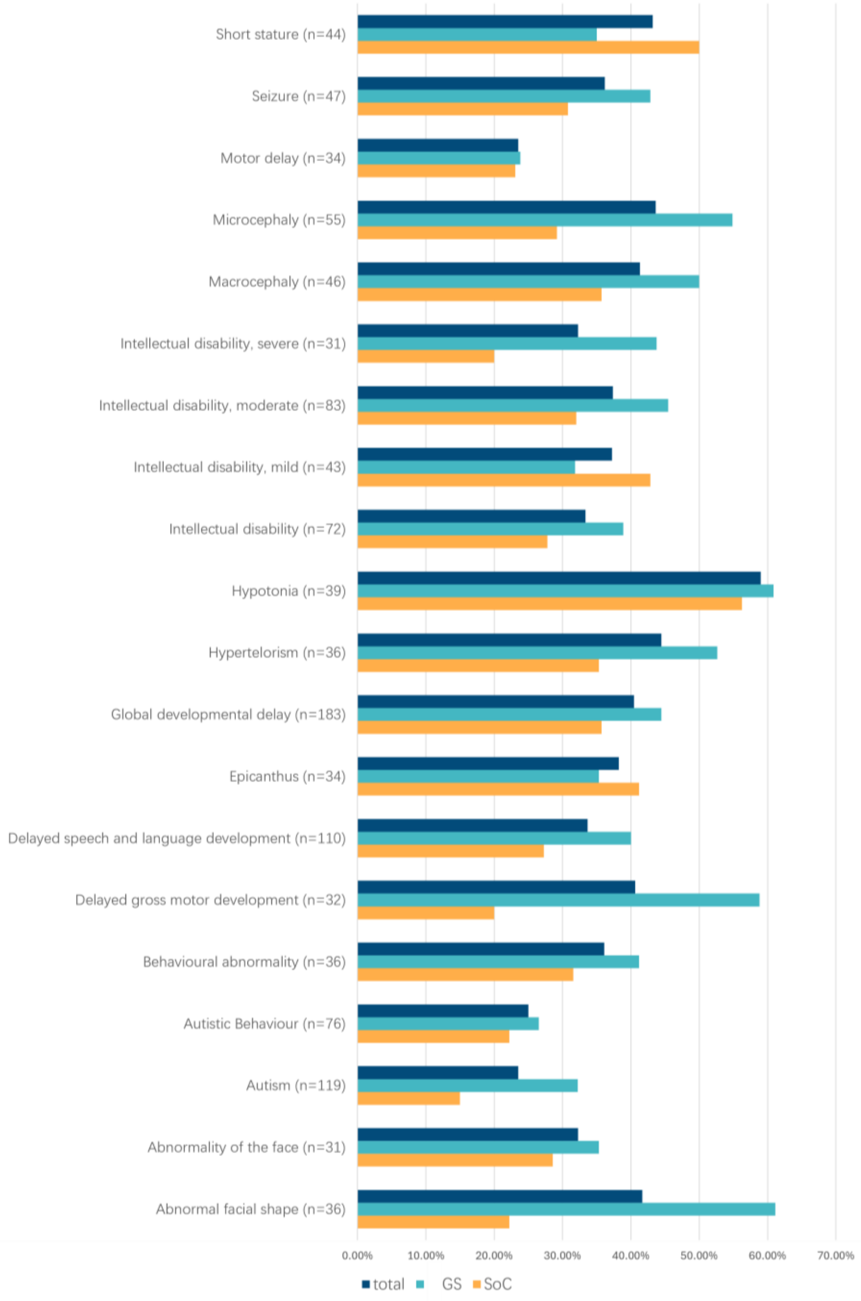


Additional Figure 6. Diagnostic yield within each study arm and across the overall cohort for the 20 most prevalent phenotypic traits, reported using the HPO terminology. The number of probands with each phenotypic trait is indicated in parentheses.

*Abbreviations:* GS, genome sequencing; HPO, human phenotype ontology; SoC, standard of care.

### **Additional Table 1:** Sequencing, bioinformatic and interpretation pipeline in each center.

### **Additional Table 1A.** Sequencing

| | *SoC - ES* | | | | | | | | | *SoC - CNVs* | | | *GS* | | | | | | |
|---|---|---|---|---|---|---|---|---|---|---|---|---|---|---|---|---|---|---|---|
| ***Center*** | ***Concentration measurment*** | ***Shearing*** | ***Shearing QC*** | ***Library preparation method*** | ***Pre-capture QC*** | ***Capture*** | ***Library QC*** | ***Sample balancing strategy*** | ***Sequencing*** | ***Technology*** | ***Platform*** | ***Resolution*** | ***Concentration measurment*** | ***Shearing*** | ***Shearing QC*** | ***Library preparation method*** | ***Library QC*** | ***Sample balancing strategy*** | ***Sequencing*** |
| ***center_1*** | Qubit | Enzymatic | - | - | - | Twist Exome | Qubit; Agilent DNA 1000 Assay (375-425bp) | - | NovaSeq 6000 | aCGH | Agilent | 60kb (in some cases 180kb) | Qubit | Tagmentation | 370-520 bp | Illumina DNA PCR free Kit | Qubit (0.8 ng/µl; >2.8 nM) | Based on volume | NovaSeq 6000 |
| ***center_2*** | Qubit | Covaris ML230 | 200-300bp | KAPA HyperPrep | - | HyperCap V3 Roche | Qubit; Fragment analyser (300-450bp) | Based on Qubit and fragment analyser results | NovaSeq 6000 | sGS | Illumina | 45kb | Qubit | Covaris ML230 | 350-450bp | KAPA Hyperprep PCR free | KAPA Library Quant kit (700-800bp) | Based on concentration and size | NovaSeq 6000 |
| ***center_3*** | Qubit | Enzymatic | - | Twist library preparation kit | Qubit | Twist Comprehensive Exome | Qubit (20-90 ng/µL); Fragment analyser (300-400bp) | Based on Qubit and fragment analyser results | NextSeq550 | aCGH | Affymetrix Cytoscan - 750K | 100kb | Outourced to center_6 | | | | | | |
| ***center_4*** | Fluoroskan | Enzymatic shearing (Twist) | 250-340bp | Twist Library Preparation EF Kit 1, 2.0 | Fluoroskan (>28ng/µl); bioanalyzer (286-390bp) | Twist Comprehensive Exome Panel | Qubit (4.6-26.8 ng/µl); Bioanalyzer (343-445bp) | Based on Qubit and fragment analyser results | NovaSeq 6000 | aCGH | Oxford Gene Technologies (OGT) Cytosure Constituional 180k v3 | Backbone 200kb; exon-level of up to 502 genes of interest | Outsourced to center_8 | | | | | | |
| ***center_5*** | Qubit | Enzymatic | - | Twist Library Preparation EF Kit | Qubit | Custom Twist Comprehensive Exome | Qubit; Tapestation | Based on Qubit and Tapestation results | NS550 or NovaSeq 6000 | SNParray/sGS | Illumina/Twist | 100kb | Outsourced to center_8 | | | | | | |
| ***center_6*** | Unchained Labs - Lunatic | Enzymatic | 250-340bp | NEBNext Ultra II FS DNA Library Prep Kit for Illumina | - | IDT xGen™ Exome Hyb Panel v2IDT | Qubit (1,5-40 ng/µl); Fragment Analyzer (280-600bp) | Based on Qubit and fragment analyser results | NovaSeq 6000 | aCGH | | 60kb | Qubit | Enzymatic | 370-520 bp | NEBNext® Ultra™ II FS DNA PCR-free Library Prep Kit | Qubit (>0,5 ng/µl); Fragment analyser (400bp) | Based on volume and corrected via spike in NovaSeq run | NovaSeq 6000 |
| ***center_7*** | Qubit | Sonication LE220 plus | 250-340bp | KAPA HyperCap | - | HyperExome V1 -KAPA HyperCAP | qPCR; Tapestation; Fluoroskan (360-480bp) | Based on qPCR concentration | NovaSeq 6000 | sGS | Illumina | 50kb | Qubit | Tagmentation | 370-520 bp | Illumina DNA PCR free Kit | Fragment analyser (420-650bp) | Based on qPCR concentration and size | NovaSeq 6000 |
| ***center_8*** | Qubit | Sonication LE220 plus | 250-340bp | KAPA HyperPrep | - | Custom Twist exome | Qubit (20-150 ng/µL); Fragment analyser (360-480bp) | Based on Qubit and fragment analyser results | NovaSeq 6000 | aCGH | Oxford Gene Technologies (OGT) Cytosure Constituional 180k v3 | Backbone 200kb; exon-level of up to 502 genes of interest | Qubit | Sonication LE220 plus | 370-520 bp | KAPA HyperPrep | Qubit (0.70-100 ng/µL); Fragment analyser (420-650bp) | Based on Qubit and fragment analyser resuts | NovaSeq 6000 |

**Additional Table 1B**. Bioinformatic pipeline

| | SoC | | | | | | | | GS | | | | | | | | |
|---|---|---|---|---|---|---|---|---|---|---|---|---|---|---|---|---|---|
| ***Center*** | ***Reference genome*** | ***Preprocessing*** | ***Mapping*** | ***Duplicate marking*** | ***Base quality score recalibration*** | ***Small variant calling*** | ***Small variant filtering*** | ***Variant score recalibration*** | ***Reference genome*** | ***Preprocessing*** | ***Mapping*** | ***Duplicate marking*** | ***Base quality score recalibration*** | ***Small variant calling*** | ***Small variant filtering*** | ***Variant score recalibration*** | ***CNV calling*** |
| ***center_1*** | hg19 | - | BWA (0.7.17) | elprep (v4.1.6) | - | GATK HaplotypeCaller & GenotypeGVCFs (3.8_1) | - | - | hg19 | - | BWA (0.7.17) | elprep (v4.1.6) | - | GATK HaplotypeCaller & GenotypeGVCFs (3.8_1) | - | GATK VariantRecalibrator & ApplyVQSR (3.8_1) | WiseCondorX |
| ***center_2*** | hg38 | - | BWA (0.7.17) | - | - | GATK HaplotypeCaller (4.2.6.1) | Hard filtering | - | hg38 | - | BWA (0.7.12) | Picard (2.18.15) | - | GATK HaplotypeCaller & GenotypeGVCFs (4.1.3.0) | - | - | QDNAseq & dnacopy |
| ***center_3*** | hg38 | - | BWA (0.7.17) | sambamba(0.7.1) | GATK BaseRecalibrator & ApplyBQSR (4.1.7) | GATK HaplotypeCaller (4.1.7) | Hard filtering | - | hg38 | - | BWA (0.7.17) | sambamba(0.7.1) | GATK BaseRecalibrator & ApplyBQSR (4.1.7) | GATK HaplotypeCaller (4.1.7) | GATK VariantFiltration (4.1.7) | - | - |
| ***center_4*** | hg38 | - | BWA (0.7.17) | Picard (2.25.5) | GATK BaseRecalibrator & ApplyBQSR (4.2.4.1) | GATK HaplotypeCaller(4.2.4.1) | - | - | hg38 | - | BWA (0.7.15) | Picard (2.18.3) | GATK BaseRecalibrator & ApplyBQSR (3.5) | GATK HaplotypeCaller (3.5) | - | GATK VariantRecalibrator & ApplyVQSR (3.5) | QDNAseq |
| ***center_5*** | hg19 | cutadapt (2.9) | BWA (0.7.17) | Picard (2.18.7) | GATK BaseRecalibrator & ApplyBQSR (4.1.3.0) | GATK HaplotypeCaller (4.1.3.0) | - | GATK VariantRecalibrator & ApplyVQSR (4.1.3.0) | hg38 | cutadapt (2.9) | BWA (0.7.17) | elprep (4.1.6) | elprep (4.1.6) | GATK HaplotypeCaller (4.1.3.0) | - | - | dnacopy |
| ***center_6*** | hg19 | - | BWA ( 0.7.10) | Picard (1.97) | GATK BaseRecalibrator & ApplyBQSR (3.3) | GATK HaplotypeCaller & UnifiedGenotyper (3.3) | Hard filtering | - | hg19 | - | BWA ( 0.7.15) | sambamba | GATK BaseRecalibrator & ApplyBQSR (4.1.5) | GATK HaplotypeCaller (4.1.5.0) | GATK VariantFiltration (4.1.5) | - | - |
| ***center_7*** | hg38 | FastP (0.23.4) | SNAP (2.0.3) | biobambam (2.0.183) | - | GATK HaplotypeCaller & GenotypeGVCFs (4.5.0.0) | - | | hg38 | - | BWA (0.7.17) | biobambam (2.0.87) | - | GATK HaplotypeCaller & GenotypeGVCFs (3.8) | - | GATK VariantRecalibrator & ApplyVQSR (3.8) | WisecondorX |
| ***center_8*** | hg38 | - | BWA (0.7.17) | Picard (2.22.1) | GATK BaseRecalibrator & ApplyBQSR (4.1.7) | GATK HaplotypeCaller & GenotypeGVCFs (4.1.7) | Hard filtering | - | hg38 | - | BWA (0.7.15) | Picard (2.16.0) | GATK BaseRecalibrator & ApplyBQSR (4.0.1.1) | GATK HaplotypeCaller (3.5) & GenotypeGVCFs (4.0.4) | GATK VariantFiltration (4.0.4) | GATK VariantRecalibrator & ApplyVQSR (4.0.4) | seqCBS (v.1.2.1) |

**Additional Table 1C.** Interpretation

| | | | center_1 | center_2 | center_3 | center_4 | center_5 | center_6 | center_7 | center_8 |
|---|---|---|---|---|---|---|---|---|---|---|
| **SoC CNV** | **Software platform(s) used for variant interpretation** | | Cartagenia Bench | In-house pipeline | In-house pipeline | OGT Cytosure Interpret | In-house pipeline | OGT Cytosure Interpret | Vivar | In-house pipeline |
| | **Reporting policy** | | class 3/4/5 | class 3+/4/5 | class 3/4/5 | class 3+/4/5 | class 3+/4/5 | class 3/4/5 | class 3/4/5 | class 3/4/5 |
| **SoC SNV** | **Software platform(s) used for variant interpretation during the study** | | Agilent Alissa Interpret | In-house pipeline | In-house pipeline | In-house pipeline (Highlander) | In-house pipeline | Highlander | In-house pipeline | In-house pipeline |
| | **Filtering strategy** | HPO-based (genome-wide) | no | yes | no | yes (exomiser filtering) | yes | no | no | no |
| | | Gene panel-based | In-house panel | Other panel | PanelApp | In-house panel | In-house panel | In-house panel | In-house panel | PanelApp |
| | **Inheritance** | *De novo* | step 1 | step 1 | step 1 | step1 | step 1 | step1 | step 1 | step 1 |
| | | X-linked | step 1 | step 2 | step 2 (males only) | step3 | step 2 | step3 | step 2 | step 2 (males only) |
| | | Autosomal recessive | step 1 | step 3 | step 3 | step2 | step 3 | step2 | step 3 | step 3 |
| | | AD inherited analysis | yes | yes | yes | yes | yes | yes | yes | yes |
| | **Workflow** | Stop analysis once pathogenic variant is detected? | no | no | yes | no | no | no | no | yes |
| | | Full exome after negative panel? | only after new request | only after new request | yes | always if trio available | only after new request | no | only after new request | yes |
| | **Reporting policy** | Primary findings | class 3+/4/5 | class 3+/4/5 | class 3/4/5 | cl3+/4/5 | class 3+/4/5 | class 3+/4/5 | class 3+/4/5 | class 3/4/5 |
| | | Secondary findings | no | opt-in for ACMG genes | no | yes with informed consent | no | no | no | no |
| **GS CNV** | **Software platform(s) used for variant interpretation** | | in-house pipeline | identical to SoC | not analyzed | Highlander | in-house pipeline | not analyzed | identical to SoC | not analyzed |
| | **Reporting policy** | | identical to SoC | identical to SoC | not applicable | not applicable | identical to SoC | not applicable | identical to SoC | not applicable |
| **GS SNV** | **Software platform(s) used for variant interpretation** | | identical to SoC | identical to SoC | identical to SoC | identical to SoC | in-house pipeline | identical to SoC | identical to SoC | identical to SoC |
| | **Filtering strategy** | HPO-based (genome-wide) | identical to SoC | identical to SoC | identical to SoC | identical to SoC | identical to SoC | identical to SoC | identical to SoC | identical to SoC |
| | | Gene panel-based | identical to SoC | identical to SoC | identical to SoC | identical to SoC | identical to SoC | identical to SoC | identical to SoC | identical to SoC |
| | **Inheritance** | *De novo* | identical to SoC | identical to SoC | identical to SoC | identical to SoC | identical to SoC | identical to SoC | identical to SoC | identical to SoC |
| | | X-linked | identical to SoC | identical to SoC | identical to SoC | identical to SoC | identical to SoC | identical to SoC | identical to SoC | identical to SoC |
| | | Autosomal recessive | identical to SoC | identical to SoC | identical to SoC | identical to SoC | identical to SoC | identical to SoC | identical to SoC | identical to SoC |
| | | AD inherited analysis | identical to SoC | identical to SoC | identical to SoC | identical to SoC | identical to SoC | identical to SoC | identical to SoC | identical to SoC |
| | **Workflow** | Stop analysis once pathogenic variant is detected? | identical to SoC | identical to SoC | identical to SoC | identical to SoC | identical to SoC | identical to SoC | identical to SoC | identical to SoC |
| | | Full exome after negative panel? | identical to SoC | identical to SoC | identical to SoC | identical to SoC | identical to SoC | identical to SoC | identical to SoC | identical to SoC |
| | **Reporting policy** | Primary findings | identical to SoC | identical to SoC | identical to SoC | identical to SoC | identical to SoC | identical to SoC | identical to SoC | identical to SoC |
| | | Secondary findings | identical to SoC | identical to SoC | identical to SoC | identical to SoC | identical to SoC | identical to SoC | identical to SoC | identical to SoC |

**Additional Table 2:** Description of demographic characteristics, inclusion criteria, phenotypes and identified molecular variants for all probands.

*cfr separate Excel table*

**Additional Table 3:** Autosomal dominant inherited variants.

- SNVs/indels

| Study ID | Study arm | variantl type | Gene | Transcript | Variant | VAF index | VAF parent | Inheritance | Phenotype carrier parent | Prior suspicion of dominantly inherited disease | Clinical relevance for the patient | Variant classification |
|---|---|---|---|---|---|---|---|---|---|---|---|---|
| center_2_001 | GS | Missense | *SCN8A* | NM_001330260.2 | c.2792G>A p.(Arg931Gln) | 0.4 | 0.45 | paternally inherited | seizures | no | Partially explains the phenotype | 3+ |
| center_2_004 | SOC | Nonsense | *TRIO* | NM_007118.4 | c.1990G>T p.(Glu664*) | 0.4 | 0.53 | maternally inherited | behavioral abnormality, intellectual disability, hyperactivity | yes | Fully explains the whole phenotype | 4 |
| center_2_028 | GS | Nonsense | *NAA15* | NM_057175.5 | c.430C>T p.(Arg144*) | 0.61 | 0.57 | maternally inherited | behavioral abnormality, autism | no | Fully explains the whole phenotype | 5 |
| center_2_060 | GS | Nonsense | *TRIO* | NM_007118.4 | c.292A>T p.(Arg98*) | 0.46 | 0.44 | maternally inherited | asymptomatic | no | Fully explains the whole phenotype | 4 |
| center_3_038 | SOC | Missense | *TSHR* | NM_000369.5 | c.122G>C p.Cys41Ser | 0.5 | 0.53 | maternally inherited | asymptomatic | no | Uncertain contribution to phenotype | 4 |
| center_5_013 | SOC | Nonsense | *EHMT1* | NM_024757.5 | c.3252C>G p.(Tyr1084*) | 0.4286 | 0.3015 | paternally inherited | normal IQ, problems with emotion regulation (tantrums) | no | Fully explains the whole phenotype | 5 |
| center_5_033 | GS | Frameshift | *POGZ* | NM_015100.4 | c.1180_1181del p.(Met394Valfs*9) | 0.609 | 0.4219 | paternally inherited | intellectual disability, special education | yes | Fully explains the whole phenotype | 5 |
| center_6_017 | SOC | Inframe indel | *ANKRD11* | NM_013275.6 | c.3835_3837del p.(Ser1279del) | 0.47 | 0.45 | maternally inherited | brachydactyly, singular crease bilateral | no | Uncertain contribution to phenotype | 3+ |
| center_6_018 | GS | Missense | *PTEN* | NM_000314.8 | c.464A>G p.(Tyr155Cys) | 0.48 | 0.43 | maternally inherited | macrocephaly, motor delay | yes | Fully explains the whole phenotype | 5 |
| center_6_020 | SOC | Inframe indel | *PTEN* | NM_000314.8 | c.160_162del p.(Val54del) | 0.34 | 0.45 | maternally inherited | motor delay, multiple thyroid nodules, breast fibroadenoma, | no | Fully explains the whole phenotype | 4 |
| center_7_014 | GS | Splice Variant | *NTRK2* | NM_006180.4 | c.213-2A>G | 0.42 | 0.49 | paternally inherited | macrocephaly, no intellectual disability | yes (partially) | Fully explains the whole phenotype | 3+ |
| center_7_017 | GS | Missense | *NSD2* | NM_133330.2 | c.3256G>A, p.(Gly1086Arg) | 0.61 | 0.02 | inherited from a mosaic parent | asymptomatic | no | Fully explains the whole phenotype | 4 |
| center_7_028 | GS | Splice Variant | *ZMYM2* | NM_197968.4 | c.3820+1G>A | 0.49 | 0.06 | inherited from a mosaic parent | asymptomatic | no | Fully explains the whole phenotype | 3+ |
| center_7_045 | GS | Frameshift | *SUFU* | NM_016169.4 | c.284dup, p.(Ser96GlufsTer2) | 0.5 | 0.51 | maternally inherited | corpus callosum agenesis, no intellectual disability | yes (partially) | Fully explains the whole phenotype | 4 |
| center_7_053 | GS | Frameshift | *DLL1* | NM_005618.4 | c.2013_2014del, p.(Glu673GlyfsTer15) | 0.56 | 0.4 | maternally inherited | asymptomatic | no | Fully explains the whole phenotype | 5 |
| center_8_037 | GS | Missense | *LZTR1* | NM_006767.3 | c.752T>G (p.Ile251Arg) | 0.446 | 0.52 | paternally inherited | asymptomatic ? pectus excavatum | no | Uncertain contribution to phenotype | 3+ |
| center_8_141 | GS | Nonsense | *GNAS* | NM_000516.7 | c.91C>T p.Gln31* | 0.469 | 0.5 | maternally inherited | short stature, one ectopic calcification | no | Fully explains the whole phenotype | 5 |
| center_8_145 | GS | Frameshift | *ZMYM2* | NM_003453.6 | c.338dupT p.Ser114Lysfs*2 | 0.495 | 0.439 | maternally inherited | difficulties with emotion regulation, autistic traits | no | Fully explains the whole phenotype | 4 |
| center_8_027 | SOC | Missense | *SYT1* | NM_005639.3 | c.845G>A p.Arg282His | 0.447 | 0.522 | maternally inherited | learning difficulties, special education | no | Fully explains the whole phenotype | 4 |
| center_8_136 | SOC | Frameshift | *TNRC6B* | NM_001162501.2 | c.2814_2815delCA (p.Ser938Argfs*9) | 0.512 | 0.402 | paternally inherited | asymptomatic ? mild learning difficulties? | no | Fully explains the whole phenotype | 4 |
| center_8_140 | SOC | Nonsense | *CACNA1C* | NM_000719_7 | c.2770C>T p.Gln924* | 0.44 | 0.2 | inherited from a mosaic parent | asymptomatic | no | Fully explains the whole phenotype | 4 |
| center_8_055 | GS | Splice variant | *DSCAM* | NM_001389.5 | c.3260-1G>A | 0.5 | 0.46 | maternally inherited | asymptomatic | yes | Uncertain contribution to phenotype | 3+ |
| center_8_163 | GS | Frameshift | *DSCAM* | NM_001389.5 | c.5274dupC (p.Ile1759Hisfs*95) | 0.471 | 0.5 | maternally inherited | special education | yes | Uncertain contribution to phenotype | 3+ |

- CNVs

| Study ID | Study arm | CNV type | CNV size (Mb) | CNV description | Inheritance | Phenotype carrier parent | Prior suspicion of dominantly inherited disease | Clinical relevance for the patient | Variant classification |
|---|---|---|---|---|---|---|---|---|---|
| center_7_022 | SOC | Loss | 0.5 | sseq[GRCh38]15q11.2(22605001_23130000)x1 | maternally inherited | mild learning disability | no | Partially explains the phenotype | 5 |
| center_7_027 | SOC | Loss | 0.3 | sseq[GRCh38] 14q24.3q31.1(78780001_79125000)x1 | paternally inherited | mild learning disability | no | Fully explains the whole phenotype | 3+ |
| center_7_030 | SOC | Loss | 0.5 | sseq[GRCh38] 15q11.2(22560001_23130000)x1 | paternally inherited | asymptomatic | no | Partially explains the phenotype | 5 |
| center_7_035 | GS | Loss | 0.5 | sseq[GRCh38] 16p12.2(21930001_22440000)x1 | maternally inherited | asymptomatic | no | Partially explains the phenotype | 5 |
| center_7_042 | GS | Gain | 2.5 | sseq[GRCh38] 22q11.21(18930001_21105000)x3 | maternally inherited | asymptomatic | no | Partially explains the phenotype | 5 |
| center_7_052 | SOC | Gain | 10 | sseq[GRCh38] 1q41q42.13(218085001_228465000)x3 | maternally inherited | short stature, mild DD | yes (partially) | Partially explains the phenotype | 3+ |
| center_7_054 | SOC | Gain | 2.4 | sseq[GRCh38] 22q11.21(19035001_21105000)x3 | paternally inherited | asymptomatic | no | Partially explains the phenotype | 5 |
| center_7_059 | GS | Gain | 1.7 | sseq[GRCh38] 1q21.1q21.2(146370001_148500000)x3 | maternally inherited | asymptomatic | no | Fully explains the whole phenotype | 5 |
| center_7_011 | GS | Loss | 0.03 | sseq[GRCh38] 16q24.3(89465001_89495000)x1 | maternally inherited | asymptomatic | no | Uncertain contribution to phenotype | 3+ |
| center_8_151 | GS | Gain | 1.98 | Seq[GRCh38] 1q21.1q21.2 (146510653_148490772)x3 | maternally inherited | asymptomatic | no | Uncertain contribution to phenotype | 5 |
| center_8_177 | GS | Gain | 1.6 | Seq[GRCh38] 7q11.23 (73195372_74773415)x3 | maternally inherited | asymptomatic | no | Fully explains the whole phenotype | 5 |
| center_8_144 | GS | Loss | 0.5 | Seq[GRCh38] 15q13.3 (31708214_32239728)x1 | paternally inherited | asymptomatic | no | Uncertain contribution to phenotype | 4 |

**Additional Table 4 :** ES coverage of SNVs/indels identified in the GS arm.

*cfr separate Excel table*

**Additional Table 5 :** Comparison of the diagnostic yield between females and males and adjusting the comparison between both study arm for these differences

- Comparison of the yields between females and males (SoC)

| dataset | female | | male | | difference | | |
|---|---|---|---|---|---|---|---|
| ± uncertain diagnoses ± AD inherited | total | diagnosed | total | diagnosed | estimate[1] | 95% CI[2] | p-value |
| certain - DN, XL, AR | 97 | 37 | 186 | 39 | 0.172 | [0.06, 0.286] | 0.002 |
| certain - DN, XL, AR, AD | 97 | 42 | 186 | 43 | 0.202 | [0.087, 0.317] | 0.001 |
| uncertain, certain - DN, XL, AR | 97 | 44 | 186 | 58 | 0.142 | [0.023, 0.261] | 0.019 |
| uncertain, certain - DN, XL, AR, AD | 97 | 51 | 186 | 62 | 0.192 | [0.071, 0.311] | 0.002 |

- Comparison of the yields between females and males (GS)

| dataset | female | | male | | difference | | |
|---|---|---|---|---|---|---|---|
| ± uncertain diagnoses ± AD inherited | total | diagnosed | total | diagnosed | estimate[1] | 95% CI[2] | p-value |
| certain - DN, XL, AR | 116 | 47 | 168 | 53 | 0.09 | [-0.024, 0.203] | 0.121 |
| certain - DN, XL, AR, AD | 116 | 55 | 168 | 58 | 0.129 | [0.013, 0.244] | 0.03 |
| uncertain, certain - DN, XL, AR | 116 | 56 | 168 | 71 | 0.06 | [-0.058, 0.177] | 0.317 |
| uncertain, certain - DN, XL, AR, AD | 116 | 67 | 168 | 80 | 0.101 | [-0.017, 0.217] | 0.093 |

- Comparison of the yields between females and males (total cohort)

| dataset | female | | male | | difference | | |
|---|---|---|---|---|---|---|---|
| ± uncertain diagnoses ± AD inherited | total | diagnosed | total | diagnosed | estimate[1] | 95% CI[2] | p-value |
| certain - DN, XL, AR | 213 | 84 | 354 | 92 | 0.134 | [0.06, 0.286] | 0.002 |
| certain - DN, XL, AR, AD | 213 | 97 | 354 | 101 | 0.17 | [0.087, 0.317] | <0.001 |
| uncertain, certain - DN, XL, AR | 213 | 100 | 354 | 129 | 0.105 | [0.023, 0.216] | 0.019 |
| uncertain, certain - DN, XL, AR, AD | 213 | 118 | 354 | 142 | 0.153 | [0.071, 0.311] | 0.002 |

- Comparison between the SoC and GS arms, adjusting for the differences between both genetic sexes

| dataset | estimate[1] | std.error | 95% CI[2] | p-value[3] |
|---|---|---|---|---|
| certain - DN, XL, AR | 0.079 | 0.038 | [0.004, 0.154] | 0.039 |
| certain - DN, XL, AR, AD | 0.09 | 0.039 | [0.013, 0.166] | 0.022 |
| uncertain, certain - DN, XL, AR | 0.082 | 0.041 | [0.001, 0.161] | 0.047 |
| uncertain, certain - DN, XL, AR, AD | 0.109 | 0.041 | [0.028, 0.190] | 0.008 |

[1] estimate of the risk difference between SoC and GS, [2] 95% confidence intervals based on Laud (2017), [3] logistic regression model

*Abbreviations:* AD, autosomal dominant inherited; AR, autosomal recessive inherited; CI, confidence interval; DN, *de novo*; GS, genome sequencing SoC, standard of care; XL, X-linked inherited.

**Additional Table 6 :** Comparison of the diagnostic yields between males and females across the different indication groups.

| Indication categories | distribution of individuals | | | | diagnostic yields (certain diagnoses) | | | |
|---|---|---|---|---|---|---|---|---|
| | male | | female | | male | | female | |
| | n= | % | n= | % | n= | % | n= | % |
| Mild to moderate ID/DD and dysmorphism | 103 | 29.10% | 74 | 34.74% | 35 | 33.98% | 34 | 45.95% |
| Mild to moderate ID/DD and family recurrence with asymptomatic parents | 27 | 7.63% | 11 | 5.16% | 6 | 22.22% | 6 | 54.55% |
| Mild to profound ID/DD and one or multiple congenital anomalies | 23 | 6.50% | 9 | 4.23% | 7 | 30.43% | 3 | 33.33% |
| Moderate to profound ID/DD | 174 | 49.15% | 94 | 44.13% | 49 | 28.16% | 45 | 47.87% |
| Multiple congenital anomalies | 11 | 3.11% | 14 | 6.57% | 1 | 9.09% | 5 | 35.71% |
| One major congenital anomaly and dysmorphism | 16 | 4.52% | 11 | 5.16% | 3 | 18.75% | 4 | 36.36% |

*Abbreviations:* ID, intellectual disability; DD, developmental delay.
